# Pseudo-neighbourhoods: Approximating the Social Characteristics of Saskatoon’s Locally-Defined Neighbourhoods using Statistics Canada’s Census Profiles

**DOI:** 10.1101/2023.08.18.23294274

**Authors:** Anousheh Marouzi, Charles Plante, Cory Neudorf

## Abstract

There is a growing desire to use social data to support local evidence-based health planning and decision-making. However, the geographic boundaries which social data are disseminated for do not usually align exactly with boundaries used by local health organizations. In this paper, we propose a method we call “pseudo-geography” to estimate counts for locally-defined geographic boundaries using data on smaller spatial units. We compared six different pseudo-geography methods, using data in Saskatoon, and identified the most accurate one, which incorporates the area-weighted spatial join technique. We further found that the pseudo-geography method can be refined by eliminating the areas with few or no residents before carrying out any spatial joins. We expect this method to be more accurate in larger cities and when the ratio of the locally-defined area to the smaller spatial units gets larger.

## Introduction

There is a growing desire to use social data to support local evidence-based health planning and decision-making (DeSalvo et al., 2016; Dubois and Lévesque, 2020). The primary source that we have for this kind of information in Canada is the Census of Population Long-form, which collects detailed information on the demographics and social-economic characteristics of approximately 25% of Canadians every five years (Statistics Canada, 2017a). However, to protect privacy, this data cannot be shared as it is directly with health organizations. Instead, Statistics Canada provides “Census Profiles” on their website, which consists of rounded summary statistics at relatively low levels of geography. Unfortunately, the geographic boundaries used by Statistics Canada for these profiles do not usually align exactly with those used by local health organizations (e.g. catchment areas for various emergency and care-providing services). Custom tabulations can be purchased from Statistics Canada for locally-defined boundaries, but this approach can be expensive and time-consuming, especially when statistics for multiple boundaries are needed.

Statistics for locally-defined geographic boundaries can be approximated by combining those provided in Statistics Canada’s Census Profiles at lower levels but the strengths and limitations of doing so, as well as how it can be done effectively, are not well understood. To support its planning and development, the City of Saskatoon routinely commissions custom tabulations of counts for multiple dozen social indicators based on its locally-defined neighbourhood boundaries. Meanwhile, the smallest level of geography for which counts are provided in Statistics Canada’s profiles is the much smaller Dissemination Area (DA). Whereas the neighbourhoods consist of approximately 4,100 persons per area, DAs consist of about 400 to 700 persons per area (Canada Statistics, 2012; City of Saskatoon Planning & Development, 2019). This provides an ideal opportunity to investigate how we can approximate the City’s “real” custom tabulation results using an approximated “pseudo-neighbourhood” estimation approach.

We considered six methods for using spatial joins to reconstitute Saskatoon’s locally-defined neighbourhoods as pseudo-neighbourhoods from DA boundaries in 2016 and used these in combination with the 2016 Census Profiles to approximate estimates of Saskatoon’s neighbourhood social characteristics. We then compared these estimates to the known amounts in the City’s custom tabulations. We were able to identify 26 indicators in the City’s data that we could recreate using our pseudo-neighbourhoods approach, including modes of travel to work, levels of education level, kinds of household composition, housing tenure, and categories of major occupation (see Appendix A for a detailed list). We used the total squared error (TSE) across all 26 indicators and neighbourhoods to compare methods.

We found that our pseudo-neighbourhoods approach was imperfect but likely sufficient for many practical applications. The most effective approximation method we used was a weighted spatial join and was, on average, accurate within about 12% across Saskatoon’s neighbourhoods; however, we also found that it could be highly inaccurate at the edges of the city where population density is more sparse and less homogenous. We also found that our approximations could be refined by considering areas known not to contain any population (e.g. water masses and the known edges of population centres.)

## Methods

### Data sources

The City of Saskatoon purchased and distributed custom tabulations (i.e., true counts) from Statistics Canada based on the 2016 census for 64 of its neighbourhoods in 2019 (City of Saskatoon Planning & Development, 2019). The 2019 version of the Saskatoon neighbourhood boundary file was retrieved from the University of Saskatchewan library and clipped only to include the 64 neighbourhoods reported by the City. Although Saskatoon consisted of as many as 96 neighbourhoods in 2019, custom tabulations were not sought for some industrial, management, and development areas (Figure 1). Three neighbourhoods, Aspen Ridge, Brighton, and U of S Lands South Management Area, were not reported on because their populations were too low at the time of the 2016 census.

**Figure 1:**
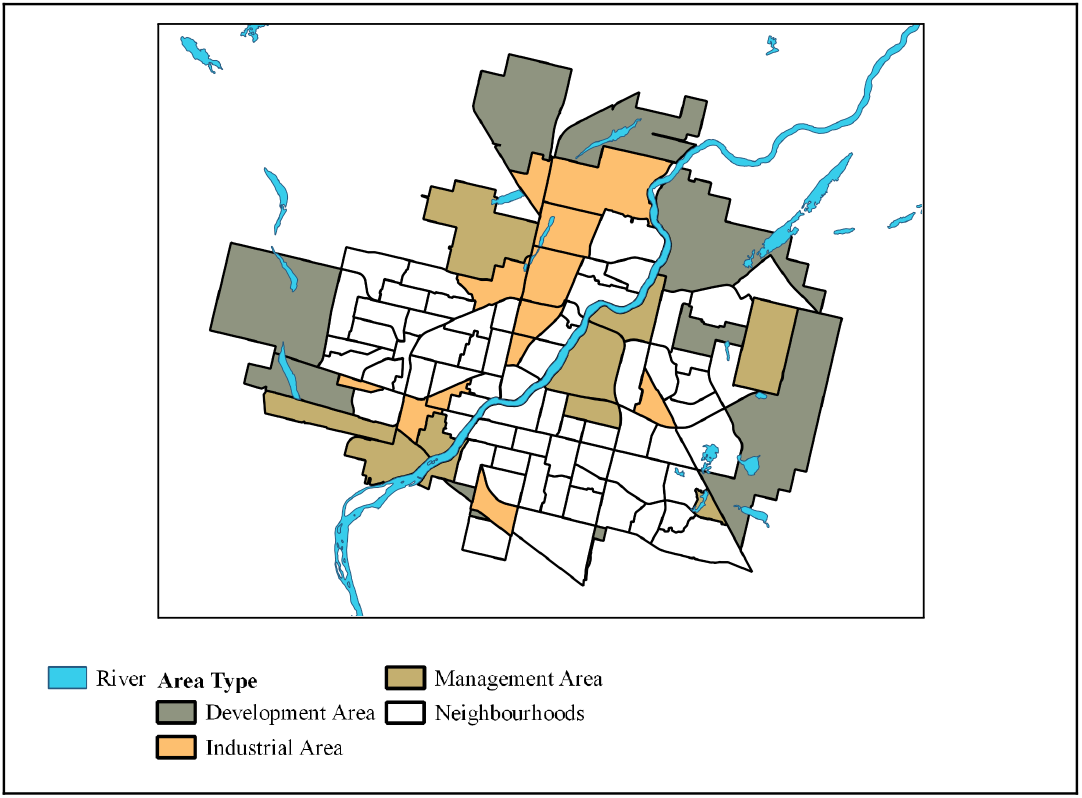
Saskatoon CSD neighbourhood boundaries (City of Saskatoon Planning and Development, 2021) Notes: The neighbourhood boundary file (2019) was requested from the University of Saskatchewan’s library.

We downloaded the Census Profile data and the nationwide DA boundary file for 2016 from Statistics Canada’s website (Statistics Canada, 2019, 2017b). These files were reduced, in turn, only to include the census subdivision (CSD) of Saskatoon. In 2016, there were 362 DAs in Saskatoon (CSD) in the Census Profile dataset and the DA boundary file. Figure 2 shows the overlap of the 362 DAs and the 64 neighbourhoods in the City of Saskatoon whose data were used in this study. We also downloaded the lake and river and the population centres boundary files used to refine our methods from Statistics Canada’s website (Statistics Canada, 2019, 2016).

**Figure 2:**
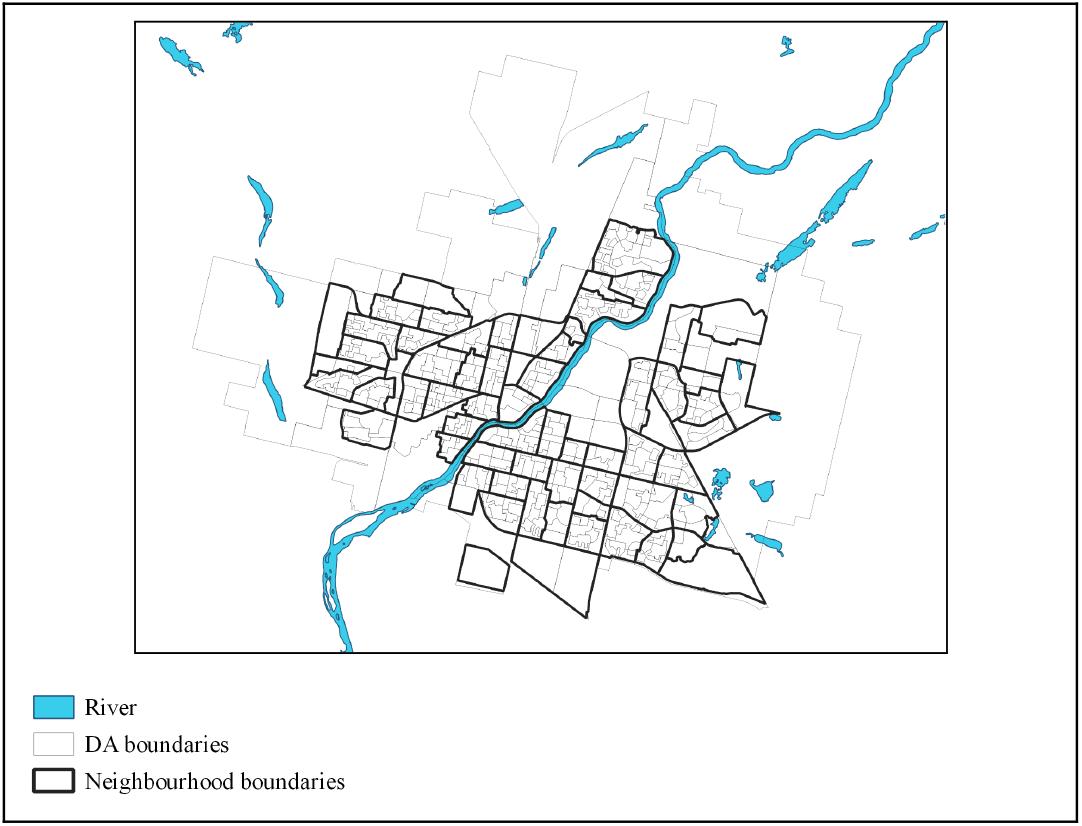
Dissemination area and neighbourhood boundaries in the city of Saskatoon Notes: The neighbourhood boundary file (2019) was requested from the University of Saskatchewan’s library. DA boundary file (2016) was derived from the Statistics Canada website.

### Constructing pseudo-neighbourhoods

Approximating true summary statistics using pseudo-geographies entails using a spatial join that joins two spatial layers based on location. A layer is the visual representation of a geographic dataset which consists of shape information (i.e., geographic boundaries or points) related to an attribute table (e.g., place names and characteristics). In our analysis, we are spatially joining Statistics Canada’s 2016 DA boundary file and Census Profiles to the City of Saskatoon’s locally-defined neighbourhood boundary file and custom tabulations. Different methods can be used to complete this spatial join, and we considered six, which we detail in this section, labelled *M*_*1*_ through *M*_*6*_. We carried out all our spatial joins using QGIS. Table 1 provides a summary of the six pseudo-neighbourhood methods in this study.

**Table 1:** Pseudo-neighbourhood methods description.

| Method | Area weighted | Get centroid of (option) | Spatial join option |
| --- | --- | --- | --- |
| M1 |  |  | contain |
| M2 |  |  | overlap |
| M3 |  | DA layer | contain |
| M4 |  | DA layer (for each part) | contain |
| M5 | Yes | Intersected layer | contain |
| M6 | Yes | Intersected layer (for each part) | contain |

Notes: M1 to M6 represents pseudo-neighbourhood methods 1 to 6.

M_1_: To create the M_1_ pseudo-neighbourhoods, we joined the DA layer’s attribute data to the locally-defined neighbourhood layer based on location, by the “contain” rule. That is, data attributed to a DA was joined to a neighbourhood if and only if no points of that DA lay outside of that neighbourhood. For example, in Figure 3, B is the only DA whose data would be joined to the neighbourhood, for the neighbourhood does not contain either A, C, or D.

**Figure 3:**
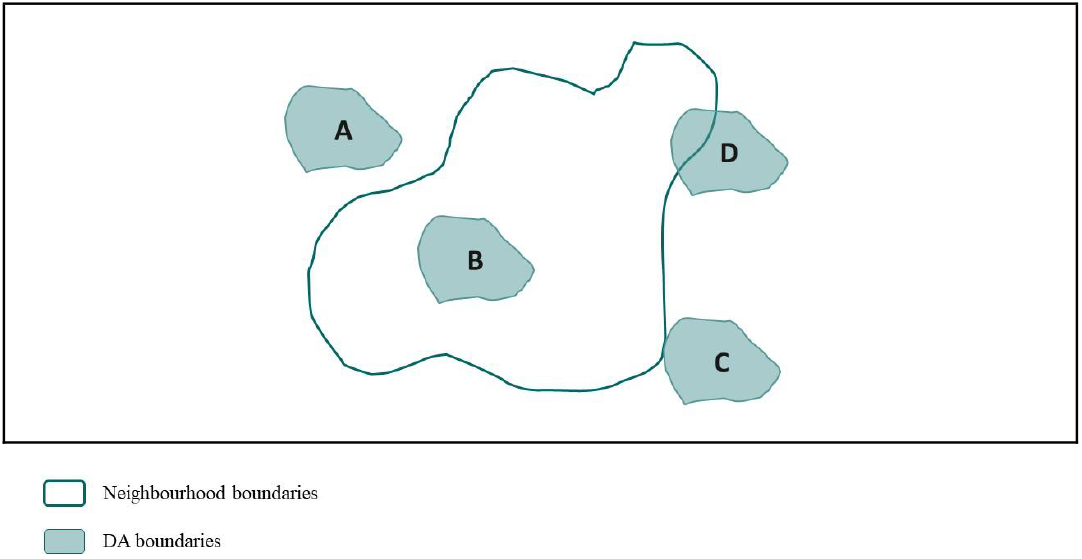
Spatial relations between layers

M_2_: To create the M_2_ pseudo-neighbourhoods, we joined the DA layer’s attribute data to the locally-defined neighbourhood layer based on location, by the “overlap” rule. In this case, data attributed to a DA was joined to a neighbourhood if the DA and neighbourhood shared space, but were not completely contained by each other. In Figure 3, only data attributed to Dissemination Area D would be joined to the neighbourhood as DAs A, B, and C do not overlap with the neighbourhood.

M_3_: To create M_3_ pseudo-neighbourhoods, we created a point layer from the DA layer, representing the centroids of each DA. We then joined this point layer and its attributed data to the neighbourhood layer to create the pseudo-neighbourhood layer, setting the join rule to “contain” (i.e., a DA would be joined to a neighbourhood if the latter contained its centroid).

M_4_: M_4_ pseudo-neighbourhoods were created in the same fashion as M_3,_ but we used a slightly different method to create the centroid layer. A centroid is a single point representing the very centre of an area. In geographic information systems, one area can be represented by two separate polygons. In such cases, the centroid of the area represents the centre of all parts of that area. To create M_4_, we stipulated that a centroid lies within each part of the shape, in cases where an area consists of more than one part.

M_5_: To further improve the accuracy of our pseudo-neighbourhood methodology, we took a different approach in the fifth attempt at creating pseudo-neighbourhoods. Before joining the DA layer to the neighbourhood layer, we calculated the proportion of each DA intersected with each locally-defined neighbourhood. This proportion was then used as a weight when joining the layers with each other based on location. In effect, we created a new weighted layer consisting of 715 areas, intersected between the DA and locally-defined neighbourhood layers. Next, we generated a centroid layer of this intersected layer, which was then spatially joined with the neighbourhood layer, setting the join rule on “contain”, (this is identical to joining without the centroid step but doing it this way builds logically to M_6_)

M_6_: M_6_ pseudo-neighbourhoods were constructed in the same way as M_5,_ but we imposed the same centroid rule used in M_4_.

### Refinement of pseudo-neighbourhood methods

After determining which pseudo-neighbourhood method was most effective at approximating true values, we considered ways for refining our results by including additional geographic information about areas that are known not to contain residents. Statistics Canada’s DA boundaries cover both land and waterbodies in Canada, whereas Saskatoon’s neighbourhoods exclude waterbodies. Before doing any spatial data manipulation, we excluded waterbodies from the DA layer so that its boundaries, specifically around the Saskatchewan River which runs through the centre of the city, align with neighbourhood boundaries. In doing so, we explored whether this difference in coverage induced bias in estimating neighbourhood-level frequencies. After excluding the waterbodies from the DA layer, we constructed the pseudo-neighbourhood layers in the same way as above. We also did the same again, this time also excluding non-population centre areas from the DA layer.

### Data Analysis

We measured the accuracy of our pseudo-neighbourhood generated counts Ĉ, by comparing them to their true counts, C, in Saskatoon’s custom tabulations. Specifically, we calculated the total square error (TSE) between them over all 26 indicators, *i*, and 64 neighbourhoods, *n*:

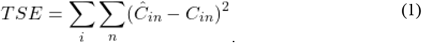

We also computed the mean relative errors (MRE) over all of the indicators for each neighbourhood separately to better understand how our approximations varied throughout the city:

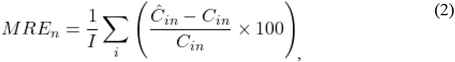

where *I* is the number of indicators.

Readers are likely to find the relative absolute errors more readily interpretable than the squared errors as they recast errors in terms of standardized units (i.e., percent of true values) although they generate substantively identical findings.

We carried out all of our computations using STATA 17.

## Results

### Comparing the pseudo-neighbourhood methods

The sixth pseudo-neighbourhood method (M_6_) was the most accurate since it had the lowest TSE compared to the other five methods. Figure 4 compares the methods’ accuracy for estimating neighbourhood statistics. Comparing M_1_ and M_2_ shows that using the contain rule spatial join (used in M_1_) was more accurate than using the overlap rule (used in M_2_). The difference between M_1_ and M_3_ shows that creating a centroid layer also improved the results of the pseudo-geography approach. The approach can be further improved by incorporating weights constructed from the intersected areas between DAs and neighbourhoods as was done in M_5_ and M_6_. We found little to no difference between M_3_ and M_4_, which means that creating the centroid layer in different ways (with or without choosing the *create centroid for each part* option) did not substantially affect the accuracy of the pseudo-neighbourhood method. Nevertheless, the marginal impact of imposing this rule between M_5_ and M_6_ was more impactful when the overall error was smaller.

**Figure 4:**
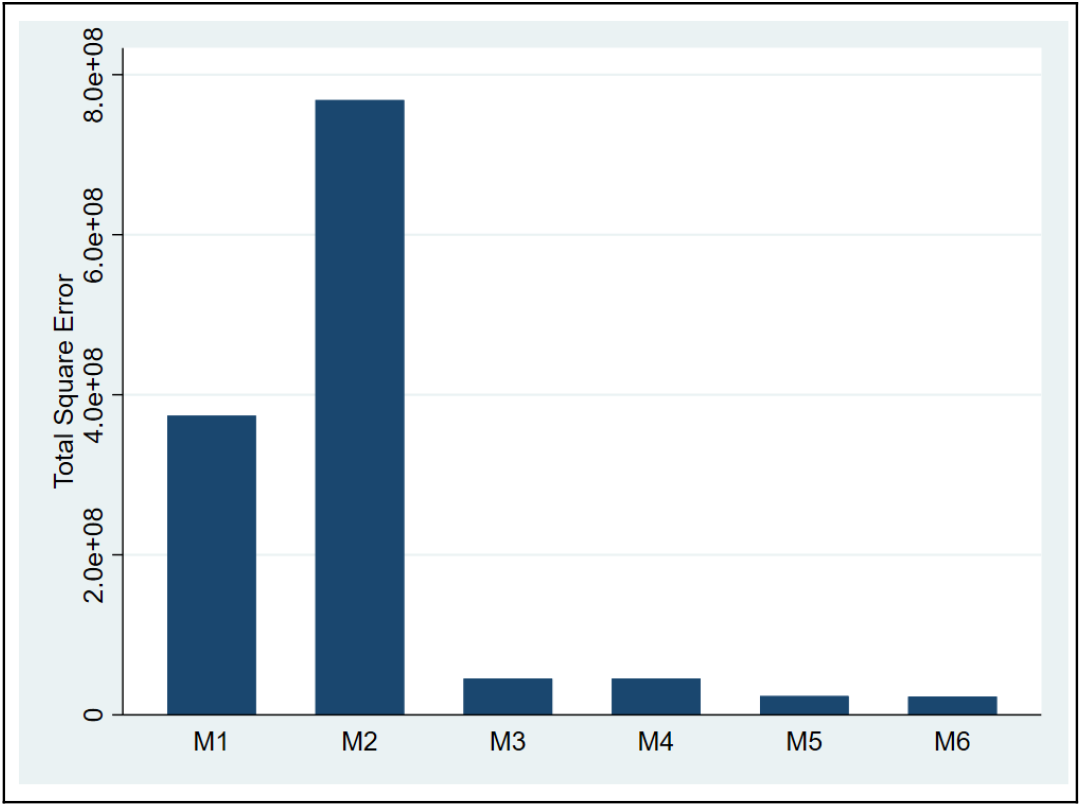
Total square error (TSE) by pseudo-neighbourhood method Notes: M_1_ to M_6_ represents pseudo-neighbourhood methods 1 to 6.

**Figure 5:**
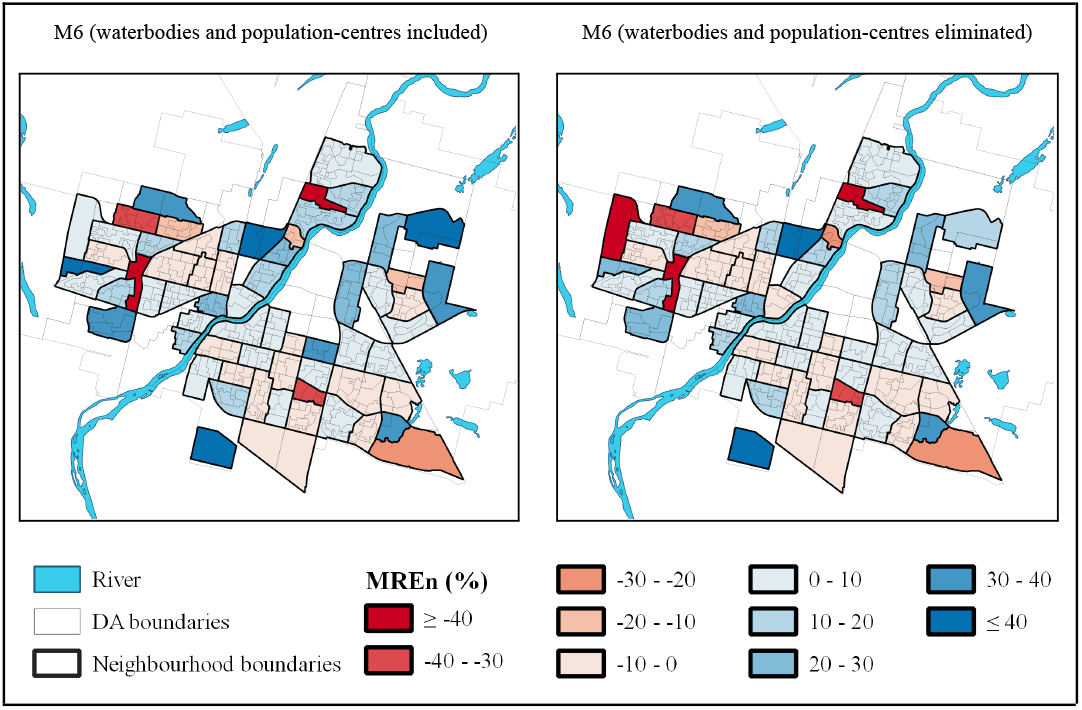
Eliminating waterbodies and non-population centres to reduce mean relative error in using pseudo-neighbourhood method Notes: Red areas (negative values) represents overcounting bias, and blue areas (positive values) represent undercounting bias. The neighbourhood boundary file (2019) was requested from the University of Saskatchewan’s library. DA boundary file (2016) was derived from the Statistics Canada website.

In addition to comparing the overall performance of the pseudo-neighbourhood methods, we also ranked them by their effectiveness at estimating each indicator. For 22 indicators, the M_6_ had the best performance and for almost all of the indicators, M_2_ was the worst. These results are consistent with what we found overall. Table 2 provides the pseudo-neighbourhood methods ranking per indicator based on the square errors calculated by indicator.

**Table 2:** Ranking pseudo-neighbourhood methods’ accuracy based on MRE_i_ per indicator.

| Variable name | Rank |  |  |  |  |  |
| --- | --- | --- | --- | --- | --- | --- |
|  | M1 | M2 | M3 | M4 | M5 | M6 |
| commute_bike | 6 | 5 | 3 | 3 | 1 | 1 |
| commute_driver | 5 | 6 | 3 | 3 | 2 | 1 |
| commute_other | 5 | 6 | 3 | 3 | 1 | 2 |
| commute_passenger | 5 | 6 | 3 | 3 | 1 | 1 |
| commute_public | 5 | 6 | 3 | 3 | 1 | 1 |
| commute_walk | 6 | 5 | 1 | 1 | 1 | 1 |
| edu_appre | 5 | 6 | 3 | 3 | 1 | 1 |
| edu_college | 5 | 6 | 3 | 3 | 2 | 1 |
| edu_highsch | 5 | 6 | 3 | 3 | 2 | 1 |
| edu_nocrt | 5 | 6 | 3 | 3 | 2 | 1 |
| hh_loneprnt | 5 | 6 | 3 | 3 | 2 | 1 |
| hh_multi | 5 | 6 | 1 | 1 | 1 | 4 |
| hh_non | 5 | 6 | 3 | 3 | 2 | 1 |
| hh_one | 5 | 6 | 3 | 3 | 2 | 1 |
| ht_owned | 5 | 6 | 3 | 3 | 2 | 1 |
| ht_rented | 5 | 6 | 3 | 3 | 2 | 1 |
| occ_art_cult_sport | 5 | 6 | 3 | 3 | 1 | 2 |
| occ_bsn | 5 | 6 | 3 | 3 | 2 | 1 |
| occ_edu_law_soci | 5 | 6 | 3 | 3 | 2 | 1 |
| occ_hlth | 5 | 6 | 3 | 3 | 2 | 1 |
| occ_manufac_uti | 5 | 6 | 3 | 3 | 1 | 1 |
| occ_mng | 5 | 6 | 3 | 3 | 2 | 1 |
| occ_nat_appsci | 5 | 6 | 3 | 3 | 2 | 1 |
| occ_natres_agr | 5 | 6 | 3 | 3 | 1 | 2 |
| occ_sale | 5 | 6 | 3 | 3 | 1 | 1 |
| occ_trd_trnsp | 5 | 6 | 3 | 3 | 2 | 1 |

Notes: Variable names are described in Appendix A; M1 to M6 represents pseudo-neighbourhood methods 1 to 6; ranking is based on the vertical comparison, that is, between different methods per variable.

The majority of the MREs by indicator for M_6_, which had the lowest TSE, were less than 10%, which is much lower than the MREs per pseudo-neighbourhood (See Appendices B and C). This conveys the fact that while the M_6_ performance may be acceptable in some pseudo-neighbourhoods, it may not be a suitable approach for estimation in other pseudo-neighbourhoods, mostly the ones that are located on the edges.

### Refining the best method

We find that by eliminating the waterbodies and non-population centres we could improve the accuracy of the pseudo-neighbourhoods estimation approach. Table 3 provides the MRE by neighbourhood for methods M_6_, M_6_ without waterbodies, and M_6_ without waterbodies and non-population centre areas. As shown in the table, the average absolute MRE_n_ for M_6_ without waterbodies (average absolute MRE_n_ = 15.85%) is lower than M_6_ (average absolute MRE_n_ = 16.46%), meaning that removing the waterbodies from the DA layer improved the pseudo-neighbourhood method. Although the average absolute MRE_n_ for M_6_ without waterbodies and non-population centre areas (average absolute MRE_n_ = 20.64%) is much higher than the average absolute MRE_n_ of M_6_ and M_6_ without waterbodies, the population-weighted average absolute MREs are not. The population-weighted average absolute MREs of M_6_ without waterbodies and non-population centre areas is only 12.09%. The primary reason for this discrepancy between unweighted and weighted results in the average absolute MREs is one new and relatively sparsely populated neighbourhood at the Western edge of Saskatoon (Kensington), for which eliminating non-population centres dramatically increased its MRE.

**Table 3:** Comparing the sixth pseudo-neighbourhood method and the ones without waterbodies and non-population centres.

| Neighbourhood | MRE<sub>n</sub> (%) |  |  | Neighbourhood | MRE<sub>n</sub> (%) |  |  |
| --- | --- | --- | --- | --- | --- | --- | --- |
|  | M6 | M6 without waterbodies | M6 without waterbodies and non-population centre areas |  | M6 | M6 without waterbodies | M6 without waterbodies and non-population centre areas |
| Adelaide/Churchill | -1 | -1 | -1 | Lakewood Urban Centre | 36 | 36 | 33 |
| Arbor Creek | 6 | 6 | 6 | Lawson Heights | 20 | 14 | 14 |
| Avalon | 15 | 15 | 11 | Lawson Heights Urban Centre | -83 | -83 | -83 |
| Blairmore Urban Centre | 56 | 56 | 21 | Massey Place | 13 | 13 | 13 |
| Brevoort Park | 6 | 6 | 6 | Mayfair | 15 | 15 | 15 |
| Briarwood | -1 | -4 | -5 | Meadowgreen | 4 | 4 | 4 |
| Buena Vista | -4 | -8 | -8 | Montgomery Place | 37 | 37 | 28 |
| Caswell Hill | 0 | 0 | 0 | Mount Royal | -8 | -8 | -8 |
| City Park | 12 | 8 | 8 | North Park | 21 | 17 | 18 |
| College Park | 2 | 2 | 2 | Nutana | 8 | 1 | 1 |
| College Park East | 1 | 1 | 1 | Nutana Park | 2 | 2 | 2 |
| Confederation Park | 5 | 5 | 5 | Nutana Urban Centre | -37 | -37 | -37 |
| Confederation Urban Centre | -76 | -76 | -75 | Pacific Heights | -4 | -4 | -4 |
| Downtown | 10 | 7 | -9 | Parkridge | 7 | 7 | 1 |
| Dundonald | -32 | -32 | -33 | Pleasant Hill | 3 | 3 | 3 |
| Eastview | -1 | -1 | -2 | Queen Elizabeth | -4 | -4 | -4 |
| Erindale | -3 | -3 | -3 | Richmond Heights | -10 | -18 | -20 |
| Evergreen | 61 | 59 | 11 | River Heights | 12 | 6 | 6 |
| Exhibition | 4 | 0 | 4 | Riversdale | 24 | 19 | 19 |
| Fairhaven | 15 | 15 | 15 | Rosewood | -22 | -22 | -24 |
| Forest Grove | 5 | 5 | 5 | Silverspring | 30 | 30 | 21 |
| Greystone Heights | 33 | 33 | 1 | Silverwood Heights | 10 | 9 | 8 |
| Grosvenor Park | -7 | -7 | -7 | Stonebridge | -2 | -2 | -2 |
| Hampton Village | 31 | 31 | 32 | Sutherland | 30 | 30 | 14 |
| Haultain | 1 | 1 | 1 | The Willows | 44 | 43 | 76 |
| Holiday Park | 14 | 9 | 8 | University Heights Urban Centre | -17 | -17 | -17 |
| Holliston | -2 | -2 | -2 | Varsity View | 9 | 9 | 8 |
| Hudson Bay Park | -7 | -7 | -7 | Westmount | -9 | -9 | -9 |
| Kelsey - Woodlawn | 44 | 44 | 41 | Westview | -19 | -19 | -20 |
| Kensington | 10 | 9 | -437 | Wildwood | 0 | 0 | 0 |
| King George | 7 | 1 | 1 | Willowgrove | 34 | 34 | 34 |
| Lakeridge | -4 | -4 | -4 | Average absolute MRE<sub>n</sub> | 16.46 | 15.85 | 20.64 |
| Lakeview | 5 | 5 | 2 | Population-weighted average absolute MRE<sub>n</sub> | 14.04 | 13.4 | 12.09 |

Notes: MRE<sub>n</sub> = mean relative error over all indicators; M6 = pseudo-neighbourhood method 6; highlighted rows indicate neighbourhoods next to the river.

Eliminating waterbodies and population centres improved the accuracy of the pseudo-neighbourhood approach and reduced the undercounting bias in many neighbourhoods, especially more densely populated neighbourhoods located along the river and at the edge of the map. Evidently, the pseudo-neighbourhood approach tended to be more accurate for more populated neighbourhoods. This is an expected side-effect of considering the MRAE which assigns greater weight to errors in less populated neighbourhoods.

## Discussion

In this paper, we propose a method to estimate counts for locally-defined geographic boundaries using boundaries and data on different and smaller spatial units, when their boundaries do not exactly align. To develop this method, we used Census DA-level data and GIS tools to generate “pseudo-neighbourhoods” and estimate Saskatoon neighbourhood-level counts for 26 indicators, in six ways using different kinds of spatial joins. We compared these six methods by their total square error and identified the most accurate methodology. We found that creating a new weighted layer consisting of intersected areas between the DA and locally-defined neighbourhood layers and then joining them using the intersected layer’s centroids was the most effective approach (i.e., M_6_). The accuracy of the pseudo-neighbourhood method is lower at the edges of cities and highest in their centres, where the population is more concentrated. Consequently, this method may work better in larger than smaller cities.

We further found that the pseudo-geography method can be refined by eliminating the areas with few or no residents, such as waterbodies and non-population centres, before carrying out any spatial joins. This method refinement is flexible, and future studies may refine their pseudo-geography approaches further by using more precise information, such as residential and commercial zoning maps.

One thing to be noted is that the amount of bias and error in the pseudo-geography approach depends on many factors, such as the range of the indicators’ frequency and the ratio of the size of the estimated area (e.g., the locally-defined neighbourhoods in this study) to the using area (e.g., the DA in this study). Considering how the pseudo-geography method works, it should become more accurate as the ratio of the estimated area to the using area increases. The locally-defined neighbourhoods used in this study are actually fairly small geographies. We anticipate that the pseudo-geography approach will perform more accurately (i.e., better than 10% error) for estimating areas with larger boundaries than the neighbourhoods, such as hospital catchment, when using DA-level data. Conversely, using this method to estimate summary statistics for smaller-boundaries areas is likely to result in greater errors.

Since we employed the pseudo-geography method in the urban context, it is hard to say how this approach might work in rural areas, and we leave consideration of this context to future studies.

## Data Availability

All data produced in the present study are available upon reasonable request to the authors.

## ^2^ Acknowledgement

The authors thank Mike Ewen, Michael Kowalchuk, and Nancy Bellegarde at the City of Saskatoon for their help with providing us with the information on Saskatoon’s custom tabulations and neighbourhood boundaries. The authors also thank Robert Alary and Sarah Rutley at the Library of the University of Saskatchewan for providing us with the boundary shapefiles. This project was funded by the Urban Public Health Network. The authors declare that they have no conflict of interest.

