## Appendices for "Pseudo-neighbourhoods: Approximating the Social Characteristics of Saskatoon’s Locally-Defined Neighbourhoods using Statistics Canada’s Census Profiles"

##### Appendix A: List of indicators used in this study.

| Variable category | Variable name | Variable description |
|---|---|---|
| Mode of travel to work | commute_bike | Bicycle |
| | commute_driver | Travel by car, truck, van (as driver) |
| | commute_other | Other types of travel |
| | commute_passenger | Travel by car, truck, van (as passenger) |
| | commute_public | Public transit |
| | commute_walk | Walking |
| Education level | edu_appre | Apprentice/trades certificate/diploma |
| | edu_college | College/CEGEP/non-university cert./dipl. |
| | edu_highsch | High school certificate or equivalent |
| | edu_nocrt | No Certificate/diploma/degree |
| Householdo structure | hh_loneprnt | Total lone-parent family households |
| | hh_multi | multiple-family households |
| | hh_non | non-family households |
| | hh_one | One-family households |
| Housing by tenure | ht_owned | Owned |
| | ht_rented | Rented |
| Major occupation | occ_trd_trnsp | Trades, transport, equip operators and related |
| | occ_art_cult_sport | Art, culture, recreation, sport |
| | occ_bsn | Business, finance, administration |
| | occ_edu_law_soci | Education, law, social, community & government |
| | occ_hlth | Health |
| | occ_manufac_uti | Manufacturing, utilities |
| | occ_mng | Management |
| | occ_nat_appsci | Natural applied sciences and related |
| | occ_natres_agr | Natural resources, agriculture & related |
| | occ_sale | Sale & service |

### Appendix B: Total counts for the city of Saskatoon (CSD) and mean relative error by indicator, data source and pseudo-neighbourhood method

| | Data Sources | | M1 | | M2 | | M3 | | M4 | | M5 | | M6 | | M6 without waterbodies | | M6 without waterbodies and non-population centre areas | |
|---|---|---|---|---|---|---|---|---|---|---|---|---|---|---|---|---|---|---|
| Variable name | Census 2016 | City of Saskatoon | TC | $MRE_i$ (%) | TC | $MRE_i$ (%) | TC | $MRE_i$ (%) | TC | $MRE_i$ (%) | TC | $MRE_i$ (%) | TC | $MRE_i$ (%) | TC | $MRE_i$ (%) | TC | $MRE_i$ (%) |
| commute_bike | 2725 | 2710 | 790 | 66 | 4520 | -90 | 2595 | 13 | 2595 | 13 | 2549 | 11 | 2566 | 9 | 2622 | 8 | 2674 | 6 |
| commute_driver | 98370 | 98095 | 23845 | 80 | 197080 | -158 | 87725 | 13 | 87725 | 13 | 86764 | 7 | 89188 | 5 | 90050 | 4 | 93999 | -3 |
| commute_other | 1835 | 1880 | 415 | 72 | 3700 | -122 | 1575 | 16 | 1575 | 16 | 1564 | 13 | 1624 | 11 | 1647 | 10 | 1716 | 7 |
| commute_passenger | 7530 | 7505 | 2075 | 78 | 14440 | -163 | 6735 | 12 | 6735 | 12 | 6670 | 6 | 6835 | 4 | 6897 | 3 | 7189 | -6 |
| commute_public | 6245 | 6170 | 1600 | 79 | 12265 | -193 | 5790 | 8 | 5790 | 8 | 5749 | 3 | 5838 | 1 | 5882 | 0 | 6068 | -10 |
| commute_walk | 5575 | 5540 | 1645 | 77 | 9965 | -174 | 5150 | -1 | 5150 | -1 | 5114 | -2 | 5150 | -3 | 5254 | -4 | 5457 | -18 |
| edu_appre | 16960 | 16925 | 4000 | 81 | 33780 | -147 | 15265 | 13 | 15265 | 13 | 15144 | 9 | 15533 | 7 | 15677 | 6 | 16354 | -5 |
| edu_college | 34045 | 33995 | 8470 | 79 | 67835 | -142 | 30775 | 14 | 30775 | 14 | 30476 | 10 | 31245 | 8 | 31563 | 7 | 32653 | 1 |
| edu_highsch | 57455 | 57230 | 14250 | 80 | 113415 | -140 | 51635 | 14 | 51635 | 14 | 51407 | 10 | 52592 | 8 | 53146 | 7 | 55060 | -1 |
| edu_nocrt | 28435 | 28350 | 7110 | 80 | 57450 | -155 | 25810 | 13 | 25810 | 13 | 25790 | 10 | 26274 | 8 | 26505 | 7 | 27388 | -5 |
| hh_loneprnt | 11020 | 11070 | 2710 | 79 | 22175 | -159 | 10020 | 11 | 10020 | 11 | 9972 | 6 | 10159 | 5 | 10243 | 4 | 10579 | -6 |
| hh_multi | 1685 | 1725 | 350 | 71 | 3670 | -128 | 1490 | 15 | 1490 | 15 | 1475 | 12 | 1532 | 9 | 1544 | 8 | 1624 | 5 |
| hh_non | 35015 | 34955 | 8055 | 82 | 72020 | -172 | 32155 | 10 | 32155 | 10 | 31756 | 8 | 32117 | 6 | 32618 | 5 | 33990 | -5 |
| hh_one | 61850 | 61685 | 15090 | 80 | 123345 | -143 | 55565 | 14 | 55565 | 14 | 55084 | 10 | 56442 | 8 | 57034 | 7 | 59135 | -1 |
| ht_owned | 65920 | 65760 | 15735 | 79 | 131775 | -137 | 59560 | 13 | 59560 | 13 | 58763 | 10 | 60286 | 8 | 60971 | 7 | 63128 | 0 |
| ht_rented | 32655 | 32525 | 7960 | 81 | 66750 | -245 | 29635 | 9 | 29635 | 9 | 29595 | 5 | 29845 | 4 | 30249 | 3 | 31636 | -24 |
| occ_art_cult_sport | 3185 | 3295 | 810 | 80 | 6075 | -154 | 2805 | 12 | 2805 | 12 | 2833 | 6 | 2895 | 4 | 2932 | 3 | 3026 | -4 |
| occ_bsn | 19890 | 19815 | 4950 | 77 | 39505 | -128 | 17870 | 16 | 17870 | 16 | 17689 | 11 | 18156 | 9 | 18350 | 8 | 19100 | 5 |
| occ_edu_law_soci | 17695 | 17655 | 4430 | 79 | 34145 | -154 | 15895 | 12 | 15895 | 12 | 15739 | 8 | 16071 | 6 | 16285 | 5 | 16883 | -2 |
| occ_hlth | 11335 | 11485 | 2730 | 80 | 22645 | -189 | 10190 | 10 | 10190 | 10 | 9956 | 3 | 10232 | 1 | 10349 | 0 | 10744 | -8 |
| occ_manufac_uti | 3860 | 3790 | 965 | 79 | 7685 | -161 | 3435 | 14 | 3435 | 14 | 3394 | 8 | 3517 | 5 | 3537 | 4 | 3719 | 0 |
| occ_mng | 13350 | 13375 | 3370 | 75 | 26205 | -156 | 11815 | 14 | 11815 | 14 | 11691 | 7 | 12016 | 5 | 12160 | 4 | 12651 | -5 |
| occ_nat_appsci | 9465 | 9385 | 2270 | 79 | 18880 | -209 | 8490 | 9 | 8490 | 9 | 8318 | 2 | 8540 | -1 | 8645 | -2 | 9012 | -12 |
| occ_natres_agr | 2730 | 2780 | 675 | 79 | 5245 | -201 | 2440 | 11 | 2440 | 11 | 2430 | 1 | 2491 | -2 | 2514 | -4 | 2582 | -9 |
| occ_sale | 33175 | 33140 | 8270 | 74 | 65965 | -135 | 29895 | 14 | 29895 | 14 | 29690 | 9 | 30376 | 8 | 30657 | 7 | 31962 | 5 |
| occ_trd_trnsp | 21785 | 21690 | 5440 | 80 | 43410 | -153 | 19385 | 13 | 19385 | 13 | 19217 | 8 | 19774 | 6 | 19942 | 5 | 20874 | -5 |

Notes: Census 2016 counts were derived from Census Profile 2016; City of saskatoon counts were derived from the City of Saskatoon's custom tabulation; TC = Total count; $MRE_i$ = mean relative error over all of the neighbourhoods; M1 to M6 represents pseudo-neighbourhood methods 1 to 6.

#### Appendix C: Total counts for the City of Saskatoon (CSD) and mean relative error by neighbourhoods and pseudo-neighbourhood method.

| | | M1 | | M2 | | M3 | | M4 | | M5 | | M6 | | M6 without waterbodies | | M6 without waterbodies and non-population centre areas | |
|---|---|---|---|---|---|---|---|---|---|---|---|---|---|---|---|---|---|
| Neighbourhood | City of Saskatoon | TC | $MRE_n$ (%) | TC | $MRE_n$ (%) | TC | $MRE_n$ (%) | TC | $MRE_n$ (%) | TC | $MRE_n$ (%) | TC | $MRE_n$ (%) | TC | $MRE_n$ (%) | TC | $MRE_n$ (%) |
| Adelaide/Churchill | 8550 | 10586 | 78 | 39090 | -282 | 7715 | 11 | 7715 | 11 | 8660 | 0 | 8681 | -1 | 8681 | -1 | 8678 | -1 |
| Arbor Creek | 10950 | 13666 | 68 | 28520 | -86 | 11345 | -6 | 11345 | -6 | 10061 | 6 | 10061 | 6 | 10061 | 6 | 10060 | 6 |
| Avalon | 8015 | 10667 | 58 | 32840 | -242 | 7130 | 16 | 7130 | 16 | 7221 | 15 | 7221 | 15 | 7227 | 15 | 7552 | 11 |
| Blairmore Urban Centre | 4360 | 1834 | 100 | 12590 | -189 | 0 | 100 | 0 | 100 | 1519 | 63 | 1831 | 56 | 1834 | 56 | 3404 | 21 |
| Brevoort Park | 8320 | 10582 | 74 | 27935 | -106 | 8325 | 5 | 8325 | 5 | 8302 | 6 | 8302 | 6 | 8302 | 6 | 8303 | 6 |
| Briarwood | 10605 | 10609 | 100 | 32155 | -181 | 6325 | 37 | 6325 | 37 | 8144 | 20 | 10249 | -1 | 10609 | -4 | 10656 | -5 |
| Buena Vista | 8230 | 10112 | 75 | 25750 | -136 | 7875 | -2 | 7875 | -2 | 8030 | -4 | 8030 | -4 | 8337 | -8 | 8335 | -8 |
| Caswell Hill | 8750 | 11240 | 70 | 25170 | -93 | 8565 | 1 | 8565 | 1 | 8666 | 0 | 8675 | 0 | 8675 | 0 | 8671 | 0 |
| City Park | 13375 | 22031 | 33 | 23565 | 9 | 12195 | 12 | 12195 | 12 | 12097 | 12 | 12097 | 12 | 12606 | 8 | 12623 | 8 |
| College Park | 12210 | 18924 | 45 | 40755 | -140 | 12070 | 2 | 12070 | 2 | 12052 | 2 | 12054 | 2 | 12054 | 2 | 12029 | 2 |
| College Park East | 10970 | 12689 | 85 | 28560 | -65 | 10990 | 0 | 10990 | 0 | 10899 | 1 | 10899 | 1 | 10899 | 1 | 10898 | 1 |
| Confederation Park | 16375 | 18210 | 89 | 37945 | -29 | 16370 | 5 | 16370 | 5 | 16312 | 5 | 16310 | 5 | 16310 | 5 | 16311 | 5 |
| Confederation Urban Centre | 2300 | 3965 | 100 | 17855 | -477 | 5415 | -140 | 5415 | -140 | 3948 | -75 | 3965 | -76 | 3965 | -76 | 3962 | -75 |
| Downtown | 7430 | 8482 | 86 | 17645 | -38 | 7420 | 6 | 7420 | 6 | 6988 | 10 | 6988 | 10 | 7347 | 7 | 8784 | -9 |
| Dundonald | 12850 | 22717 | 50 | 48980 | -234 | 8325 | 36 | 8325 | 36 | 11481 | 10 | 16292 | -32 | 16292 | -32 | 16458 | -33 |
| Eastview | 8675 | 11501 | 74 | 38470 | -234 | 8385 | 10 | 8385 | 10 | 9396 | -1 | 9396 | -1 | 9396 | -1 | 9414 | -2 |
| Erindale | 9515 | 10907 | 88 | 28560 | -100 | 10760 | -13 | 10760 | -13 | 9593 | -2 | 9692 | -3 | 9692 | -3 | 9692 | -3 |
| Evergreen | 13100 | 5232 | 100 | 30795 | -140 | 0 | 100 | 0 | 100 | 5055 | 61 | 5055 | 61 | 5232 | 59 | 11285 | 11 |
| Exhibition | 7685 | 7756 | 100 | 23530 | -115 | 6770 | 13 | 6770 | 13 | 6980 | 9 | 7382 | 4 | 7756 | 0 | 7392 | 4 |
| Fairhaven | 11305 | 11341 | 82 | 26430 | -68 | 8545 | 26 | 8545 | 26 | 9611 | 15 | 9611 | 15 | 9611 | 15 | 9610 | 15 |
| Forest Grove | 14175 | 19475 | 58 | 33980 | -56 | 13385 | 5 | 13385 | 5 | 13389 | 5 | 13455 | 5 | 13455 | 5 | 13451 | 5 |
| Greystone Heights | 5650 | 4012 | 100 | 17305 | -141 | 3565 | 41 | 3565 | 41 | 4009 | 33 | 4012 | 33 | 4012 | 33 | 5673 | 1 |
| Grosvenor Park | 3845 | 3872 | 100 | 15255 | -233 | 3490 | 8 | 3490 | 8 | 3872 | -7 | 3872 | -7 | 3872 | -7 | 3873 | -7 |
| Hampton Village | 19215 | 13217 | 100 | 51570 | -42 | 24230 | -26 | 24230 | -26 | 13216 | 31 | 13217 | 31 | 13217 | 31 | 13177 | 32 |
| Haultain | 7860 | 10698 | 69 | 21130 | -78 | 8050 | 4 | 8050 | 4 | 7998 | 1 | 7998 | 1 | 7998 | 1 | 7997 | 1 |
| Holiday Park | 3860 | 2959 | 100 | 10335 | -134 | 2715 | 16 | 2715 | 16 | 2825 | 14 | 2825 | 14 | 2959 | 9 | 3035 | 8 |
| Holliston | 8795 | 13340 | 51 | 26165 | -83 | 9450 | -9 | 9450 | -9 | 9000 | -2 | 9000 | -2 | 9000 | -2 | 9000 | -2 |
| Hudson Bay Park | 5080 | 6575 | 69 | 22550 | -236 | 6280 | -30 | 6280 | -30 | 5165 | -6 | 5185 | -7 | 5185 | -7 | 5185 | -7 |
| Kelsey - Woodlawn | 2400 | 1517 | 100 | 13025 | -446 | 1120 | 63 | 1120 | 63 | 1509 | 44 | 1516 | 44 | 1517 | 44 | 1584 | 41 |
| Kensington | 1640 | 1388 | 100 | 16550 | -1024 | 0 | 100 | 0 | 100 | 1219 | 19 | 1373 | 10 | 1388 | 9 | 7283 | -437 |
| King George | 4725 | 4356 | 100 | 15440 | -144 | 4725 | -7 | 4725 | -7 | 4124 | 7 | 4124 | 7 | 4356 | 1 | 4357 | 1 |

| | | | | | | | | | | | | | | | | | |
|---|---|---|---|---|---|---|---|---|---|---|---|---|---|---|---|---|---|
| Lakeridge | 8660 | 11775 | 62 | 32965 | -298 | 8855 | -10 | 8855 | -10 | 8369 | -4 | 8370 | -4 | 8370 | -4 | 8368 | -4 |
| Lakeview | 17065 | 25908 | 42 | 29650 | 14 | 15120 | 11 | 15120 | 11 | 16118 | 5 | 16118 | 5 | 16118 | 5 | 16602 | 2 |
| Lakewood Urban Centre | 5955 | 3672 | 100 | 22175 | -258 | 1635 | 70 | 1635 | 70 | 3653 | 36 | 3669 | 36 | 3672 | 36 | 3871 | 33 |
| Lawson Heights | 11630 | 12003 | 85 | 31000 | -80 | 11055 | 3 | 11055 | 3 | 9315 | 20 | 9315 | 20 | 9898 | 14 | 9862 | 14 |
| Lawson Heights Urban Centre | 4205 | 6377 | 100 | 20245 | -415 | 4900 | -24 | 4900 | -24 | 6377 | -83 | 6377 | -83 | 6377 | -83 | 6377 | -83 |
| Massey Place | 7620 | 7802 | 88 | 16045 | -37 | 5760 | 29 | 5760 | 29 | 6862 | 13 | 6862 | 13 | 6862 | 13 | 6861 | 13 |
| Mayfair | 6795 | 6911 | 85 | 17010 | -77 | 5705 | 16 | 5705 | 16 | 5712 | 15 | 5714 | 15 | 5716 | 15 | 5717 | 15 |
| Meadowgreen | 10050 | 13176 | 63 | 23460 | -48 | 9360 | 7 | 9360 | 7 | 9516 | 5 | 9556 | 4 | 9556 | 4 | 9554 | 4 |
| Montgomery Place | 7255 | 5818 | 83 | 17600 | -147 | 2515 | 66 | 2515 | 66 | 4503 | 37 | 4503 | 37 | 4503 | 37 | 5190 | 28 |
| Mount Royal | 11285 | 16513 | 51 | 30385 | -93 | 11460 | -12 | 11460 | -12 | 11183 | -8 | 11183 | -8 | 11183 | -8 | 11185 | -8 |
| North Park | 5340 | 5711 | 80 | 14380 | -81 | 5320 | 4 | 5320 | 4 | 4197 | 23 | 4295 | 21 | 4496 | 17 | 4478 | 18 |
| Nutana | 17180 | 22522 | 64 | 40475 | -49 | 15805 | 6 | 15805 | 6 | 15359 | 8 | 15359 | 8 | 16667 | 1 | 16695 | 1 |
| Nutana Park | 6710 | 6386 | 100 | 47125 | -555 | 3915 | 41 | 3915 | 41 | 6391 | 2 | 6386 | 2 | 6386 | 2 | 6392 | 2 |
| Nutana Urban Centre | 6150 | 5326 | 100 | 17575 | -300 | 5425 | -37 | 5425 | -37 | 5326 | -37 | 5326 | -37 | 5326 | -37 | 5327 | -37 |
| Pacific Heights | 9885 | 15558 | 41 | 35630 | -169 | 10030 | -4 | 10030 | -4 | 9949 | -3 | 9993 | -4 | 9993 | -4 | 9997 | -4 |
| Parkridge | 11190 | 14912 | 64 | 34720 | -124 | 10750 | 6 | 10750 | 6 | 9393 | 18 | 10670 | 7 | 10672 | 7 | 11251 | 1 |
| Pleasant Hill | 8520 | 9753 | 89 | 21065 | -38 | 9050 | -2 | 9050 | -2 | 8718 | 3 | 8718 | 3 | 8718 | 3 | 8719 | 3 |
| Queen Elizabeth | 7120 | 7131 | 100 | 20905 | -95 | 7205 | -8 | 7205 | -8 | 7127 | -4 | 7131 | -4 | 7131 | -4 | 7131 | -4 |
| Richmond Heights | 1870 | 2026 | 100 | 7770 | -323 | 1015 | 45 | 1015 | 45 | 1899 | -10 | 1900 | -10 | 2026 | -18 | 2045 | -20 |
| River Heights | 10405 | 11885 | 78 | 23600 | -30 | 10300 | 2 | 10300 | 2 | 9184 | 12 | 9214 | 12 | 9860 | 6 | 9871 | 6 |
| Riversdale | 4865 | 3950 | 100 | 14835 | -142 | 3545 | 30 | 3545 | 30 | 3751 | 24 | 3751 | 24 | 3950 | 19 | 3983 | 19 |
| Rosewood | 7995 | 9028 | 100 | 32150 | -163 | 12285 | -66 | 12285 | -66 | 9027 | -22 | 9027 | -22 | 9028 | -22 | 9175 | -24 |
| Silverspring | 12260 | 11069 | 84 | 31180 | -118 | 6720 | 48 | 6720 | 48 | 8191 | 36 | 8807 | 30 | 8874 | 30 | 9883 | 21 |
| Silverwood Heights | 24430 | 34375 | 51 | 45695 | 5 | 22725 | 8 | 22725 | 8 | 21957 | 11 | 22129 | 10 | 22565 | 9 | 22689 | 8 |
| Stonebridge | 25935 | 25920 | 100 | 65830 | -40 | 30435 | -20 | 30435 | -20 | 25920 | -2 | 25920 | -2 | 25920 | -2 | 26036 | -2 |
| Sutherland | 14405 | 19353 | 39 | 21765 | 7 | 9050 | 39 | 9050 | 39 | 10273 | 30 | 10273 | 30 | 10303 | 30 | 12437 | 14 |
| The Willows | 1685 | 943 | 100 | 0 | 100 | 0 | 100 | 0 | 100 | 916 | 44 | 916 | 44 | 943 | 43 | 395 | 76 |
| University Heights Urban Centre | 4245 | 4133 | 100 | 34225 | -901 | 1435 | 61 | 1435 | 61 | 4102 | -16 | 4127 | -17 | 4133 | -17 | 4138 | -17 |
| Varsity View | 9690 | 11362 | 75 | 24850 | -74 | 8975 | 5 | 8975 | 5 | 8830 | 9 | 8830 | 9 | 8837 | 9 | 8859 | 8 |
| Westmount | 6350 | 8314 | 76 | 22260 | -179 | 6515 | 2 | 6515 | 2 | 6929 | -9 | 6944 | -9 | 6944 | -9 | 6943 | -9 |
| Westview | 8060 | 11236 | 71 | 27950 | -210 | 4520 | 42 | 4520 | 42 | 7000 | 9 | 9096 | -19 | 9096 | -19 | 9133 | -20 |
| Wildwood | 19735 | 25858 | 67 | 46890 | -35 | 19775 | 0 | 19775 | 0 | 19920 | 0 | 19927 | 0 | 19933 | 0 | 19931 | 0 |
| Willowgrove | 17170 | 14123 | 84 | 45790 | -45 | 21005 | -21 | 21005 | -21 | 11524 | 34 | 11524 | 34 | 11378 | 34 | 11373 | 34 |

Notes: City of saskatoon counts were derived from the City of Saskatoon's custom tabulation; TC = Total count; $MRE_n$ = mean relative error over all of the indicators; M1 to M6 represents pseudo-neighbourhood methods 1 to 6.
